# Field evaluation of Rapid SARS-Cov2 Antigen screening test on self-collected deep throat saliva samples in Malaysia

**DOI:** 10.1101/2021.12.20.21268141

**Authors:** Noorliza Mohamad Noordin, Steven Chee Loon Lim, Zhuo-zhi Lim, Teck-Onn Lim, On behalf of the Covid19 Screening Study group

## Abstract

Low cost Rapid Antigen Tests are widely used in Malaysia and the government has also mandated worksite screening as a condition for reopening. Numerous RAT kits have been approved by the Malaysian Medical Device Authority. However, it remains uncertain how these kits would perform in the field.

We enrolled workers between June and September 2021 from 23 worksites. They were trained and experienced in performing RAT selftest by virtue of their worksite participation in routine screening program. These workers also had reverse transcriptase polymerase chain reaction tests in the course of mass screening or contact tracing. We also enrolled patients with PCR confirmed Covid19 from a quarantine centre. These patients were instructed on selftesting and then immediately perform RAT under supervision. Two manufacturers donated RAT for this study.

A total of 340 participants were enrolled, 130 were from quarantine centre and 210 from worksites. The overall sensitivity of RAT compared to PCR was 70 percent. The specificity was 91 percent. Sensitivity decreased with increasing PCR cycle threshold values. Sensitivity is also lower among untrained subjects at each level of Ct. Logistic regression analysis confirmed false negative result is associated with Ct and participants prior training and experience.

This study shows that in the real world, RAT performance were markedly lower than that reported by the manufacturers. The test sensitivity is dependent on the operator training and experience, as well as on viral load as measured by Ct. User training and repeated testing for screening purpose is necessary to mitigate the low sensitivity of RAT.

## Introduction

Malaysia has among the highest Covid19 incidence and mortality in Asia^1^. The government has ramped up vaccination rapidly, reaching almost 100% adult coverage by October 2021, which has enabled the country to transition to the endemic phase. With nationwide lockdown lifted, Malaysia’s control strategy has shifted accordingly to minimizing hospitalizations and deaths. Self-testing is widely deployed as a control measure, among others. This is to enable early detection and self-isolation. Rapid Antigen Test (RAT) based on lateral flow assay for detection of SARS-Cov2 antigen^2^ on self-collected deep throat saliva sample is recommended^3,4^. RAT is simple, safe, affordable and takes <20 minutes to produce results. To encourage its uptake, low cost (∼ USD 1.2 per kit) RAT is widely available at pharmacies and supermarkets. The government has also mandated worksite screening as a condition for reopening^5^. This is because Malaysia hosted a large number of migrant workers (22% of our labour force^6^) and Covid19 outbreaks had frequently occurred at worksites (factories, construction sites, farms, plantations) where large number of migrant workers are found working in closed setting and living in crowded dormitories.

Numerous RAT kits have been approved by the Malaysian Medical Device Authority (MDA)^7^. All kits have been independently validated by MDA-designated labs, and all reported sensitivity of 95% and specificity 99% or better. However, it remains uncertain how these kits would perform in the field where individuals perform self-test at homes, worksites and other congregate settings.

## Methods

We conducted a cross sectional study to evaluate the diagnostic performance of RAT for detecting SARS-Cov2 antigen. The Ministry of Health’s (MOH) Medical and Research Ethics Committee approved the study and all subjects gave written informed consent.

We enrolled workers between June and September 2021 from 23 worksites. They were trained and experienced in performing RAT self-test by virtue of their worksites’ participation in routine screening program. These workers also had reverse transcriptase polymerase chain reaction (RT-PCR) tests in the course of mass screening or contact tracing. Each subject was tested once only using the RAT deployed at the worksite.

We further enrolled another 130 subjects with RT-PCR confirmed Covid19 who were admitted into a quarantine centre (Pusat Kuarantin & Rawatan Covid19 Berisiko Rendah Ipoh PKRC). These subjects were given instructional material and video to read and watch. They also received brief instruction on collecting deep throat saliva and on the test procedure. The subjects then immediately collect their own saliva samples and perform the RAT, under the supervision of trained health professionals. Each subject was tested twice using 2 brands of RAT (Ecotest, Getein). Manufacturers of these RAT have agreed to contribute their kits for this evaluation, while others invited have declined. All tests were performed according to the manufacturers’ instructions. In the event of invalid result, the test was repeated until a valid result was obtained.

## Results

A total of 340 participants were enrolled, 130 were from quarantine centre and 210 from worksites. Their mean age were 31 years, 76% were male and 53% were migrant workers (Table 1), reflecting the selection of worksites for this study. The overall sensitivity (Table 2) of RAT compared to RT-PCR was 70% (95% CI 65%-75%). The specificity was 91% (95% CI 84%-95%).

**Table 1:** Baseline characteristics of study participants.

|  | Quarantine centre | Worksites | All Subjects |
| --- | --- | --- | --- |
| Total number | 130 | 210 | 340 |
| **Demographic, No. (%)** |  |  |  |
| Age, year |  |  |  |
| Mean (SD) | 30 (10) | 32 (8) | 31 (9) |
| Median (IQR) | 29 (22,37) | 30 (26,36) | 29 (25,36) |
| Gender, No. (%) |  |  |  |
| Male | 68 (52%) | 190 (90%) | 258 (76%) |
| Female | 62 (48%) | 20 (10%) | 82 (24%) |
| Ethnicity, No. (%) |  |  |  |
| Malay | 112 (86%) | 16 (8%) | 128 (38%) |
| Chinese | 3 (2%) | 9 (4%) | 12 (3%) |
| Indian | 15 (12%) | 4 (2%) | 19 (6%) |
| Migrant workers | 0 (0%) | 181 (86%) | 181 (53%) |

**Table 2:** Overall diagnostic performance of Rapid test for SARS-Cov2 antigen compared to RT-PCR test as reference.

|  | RT-PCR negative | RT-PCR Positive |
| --- | --- | --- |
| RAT Positive | 14 | 224 |
| RAT Negative | 137 | 95 |
| Total tests | 151 | 319 |
|  | Overall Specificity (95% CI)<br>91% (84%, 95%) | Overall Sensitivity (95% CI)<br>70% (65%, 75%) |

Sensitivity decreased with increasing PCR cycle threshold (Ct) values from 87% in samples with a Ct of <20 to 18% in samples with Ct value ≥30. Sensitivity also increased with participants’ prior training and experience. Test sensitivity is lower among untrained subjects at each level of Ct value (Table 3)

**Table 3:** Sensitivity of Rapid test for SARS-Cov2 antigen compared to RT-PCR test as reference in relation to PCR cycle threshold value and Participants’ prior training and experience.

|  |  | All participants | Trained and experienced participants | Untrained and inexperienced participants |
| --- | --- | --- | --- | --- |
|  | No. (%) | 334 | 59 | 260 |
| **PCR cycle threshold (Ct) values** | No. (%) |  |  |  |
| <20 | 168 | 87% (81,92) | 98% (88,100) | 83% (75,89) |
| 20- <30 | 113 | 63% (53,72) | 91% (59,100) | 60% (50, 69) |
| >=30 | 38 | 18% (8,34) | 50% (7,93) | 15% (5,31) |

Multivariate logistic regression analysis confirmed that false negative result (1-sensitivity) is strongly associated with Ct value (OR=1.18, 95% CI 1.13-1.23, p<0.001) and participants’ prior training and experience (OR=0.18, 95% CI 0.00-0.04, p<0.001). Each unit increase in Ct value increases the risk of false negative result by 18%, while training decreases risk by 82%.

## Discussion

This study shows that in the field, test performance of RAT differs markedly from lab based validation reported by the manufacturers and MDA. In particular, the specificity was lower than the reported 99%. This is consistent with frequent observation of false positive results in the screening context where the pre-test probability is low. The high risk of false positive results is of concern, which has previously led to regulatory authority recalling such kits^8^.

Our results also show the test sensitivity is dependent on the operators’ training and experience, as well as on viral load as measured by Ct, consistent with other report^9,10,11^. At Ct value<20, the sensitivity was 87% which decreased to only 18% at Ct>30. This was lower than the 95% or better reported by the manufacturers and MDA. RAT based screening will often produce false negative results especially when performed by untrained individuals and early in SARS-CoV-2 infection when the viral load is low, albeit at this stage of the infection, these individuals are not highly infectious. This underscores the importance of users’ training and frequent repeat testing in the screening context to mitigate the low sensitivity of RAT^12^.

This study has limitations. The sample size is small. While we have used a convenient sample, deliberately enriched with RT-PCR positive subjects, the subjects are likely to be representative of the population who routinely undergo screening in Malaysia. We have also not been able to collect data on symptoms, another factor known to affect test performance^9,10,11^. Some RT-PCR testing were performed a day or 2 prior to RAT, which may also influence the results.

## Data Availability

All data produced in the present study are available upon reasonable request to the authors

